# Prediction of Peak and Termination of Novel Coronavirus Covid-19 Epidemic in Iran

**DOI:** 10.1101/2020.03.29.20046532

**Authors:** Amir-Pouyan Zahiri, Sepehr RafieeNasab, Ehsan Roohi

## Abstract

The growth and development of Covid-19 transmission have significantly cut the attention of many societies, particularly Iran that has been struggling with this contagious, infectious disease since late February 2020. In the present study, the known SIR model was used for the dynamics of an epidemic to provide a suitable assessment of the COVID-19 virus epidemic in Iran. The epidemic curve and SIR model parameters were obtained with the use of Iran statistical data. The recovered people were considered alongside the official number of confirmed victims as the reliable long-time statistical data of Iran. The results offered many important predictions of the COVID-19 virus epidemic such as realistic number of victims, infection rate, peak time, and other characteristics.

## Introduction

The 2019 coronavirus (COVID-19) pandemic is a global outbreak of coronavirus disease. Given the economic and social impacts of this phenomenon, mathematical modeling and prediction are essential to aware people and decision-making managers of the consequences of this epidemic. Accordingly, the present research team has mathematically investigated the COVID-19 growth in Iran using the available statistical data and epidemic curve. Available official long-time data of Iran were used to predict the number of victims (V) of this virus in this country. Different types of mathematical models can be used for predicting an epidemic infection, and many important unknown variables can be achieved by the use of real available data. The first predictions of an epidemic infection exponential growth, that is typical for the initial stages of all epidemics, have been presented in [1].

More complicated mathematical models were obtained as the susceptible-exposed-infectious-recovered (SEIR) model. SEIR model is engaged upon anticipating the potential domestic and international spread of the COVID-19 outbreak [2]. The intricate models need more attempts for the man parameter identification. Accordingly, Nesteruk [3, 4] developed an appropriate mathematical model of contamination and SIR-model of spreading an infection to predict the time dynamics of the unknown children’s disease, which occurred in Chernivtsi (Ukraine), and COVID-19 China epidemic. In this paper, the known SIR model for the dynamics of an epidemic [5-8] is used to identify Iran (COVID-19) epidemic in Iran. In this respect, an exact solution of the SIR linear equations is considered simultaneously with a statistical approach based on the confirmed and recovered victims as the reliable long-time statistical data of Iran [4]. Main epidemic characteristics such as epidemic victim numbers, infected, and recovered people are estimated over time. Furthermore, Iran’s official data [9, 10] are fitted to the China epidemic curve [11-13] and compared with the results of the SIR model.

- STEP 1

### Exact solution of SIR-equations

The SIR-model linear equations are illustrated for an infectious disease [3, 4], as below:

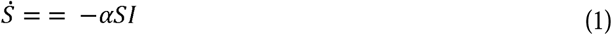

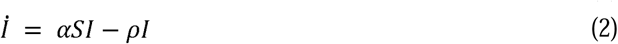

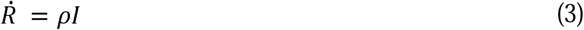

The number of infected people is denoted by *I*, susceptible by S, recovered by *R*. Also, the immunization and infection rates are represented by *ρ* and α, respectively. Given that 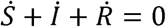 (Eqs. (1-3)), *N = S + I + R* needs to be unchanged over time and can be dealt with as the number of susceptible people before the epidemic outbreak because of *I* = *R* = 0 at *t* < *t*_0_. It is worth noting that *N* is not equal to the total population (*N*_*total*_), but rather it stands for the initial number of sensitive people who are unprotectively prone to some special diseases. Particularly, *N*/*N*_*total*_ might be quite small. For instance, there are 3711 people on the board of Diamond Princess, with confirmed cases of 70 on 10.02.2020, i.e., the susceptible people can be estimated by 1.89%.

Relation (4) is supposed to define the initial conditions for Eqs. (1-3).

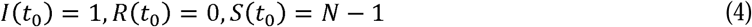

It follows from Eqs. (1) and (2):

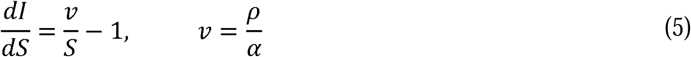

Integration of Eq. (5) with the initial conditions presented in relation (4) results in:

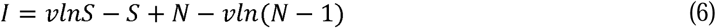

Function *I* reaches its maximum value at *S* = *ν* and approaches zero-value at infinity. Also, the number of susceptible people at infinity (*S*_∞_> 0) can be calculated via Eq. (6).

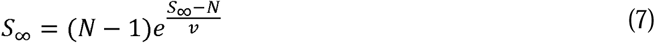

Eqs. (1-3) were solved by proposing *V*(*t*) = *I*(*t*) + *R*(*t*), corresponding to the number of victims. The corresponding equation integration would be [3]:

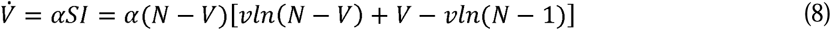

Resulting in:

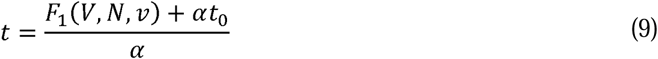

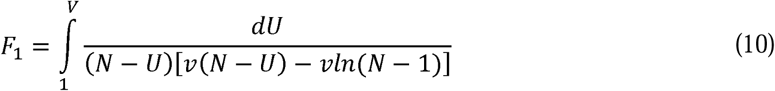

Therefore, for every set of *N, v, α, t*_0_ and a constant value of *V*, Eq. (10) can be solved, and the corresponding time can be calculated using Eq. (9). Then, *I* can be calculated using Eq. (6) by replacing *S* = *N* − *V* and *R* out of *R* = *V* − *I*.

Also for more comprehensible and convenient prediction, can be used two approximations to calculate random function *F*_1_(*V, N, ν*) and time function (t), subsequently [3].

The obtained solution of the differential equations (1-3) can be simplified with the use of these different approximations for the function ln(*N* − *U*) − ln (*n* − 1).

If we assume *ln*(*N* − *U*) − *ln* (*n* − 1) ≈ 0 then 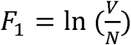 and ultimately:

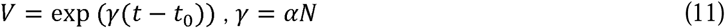

According to (11) epidemic starts exponentially and only two parameters (*γ and t*_0_) describe the process.

If we would like to anticipate more appropriately and accurately, we can assume ln(*N* − *U*) − ln (*N* − 1) ≈ (*1* −*U*)/*N*, then 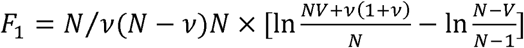.

And the solution’s form will be:

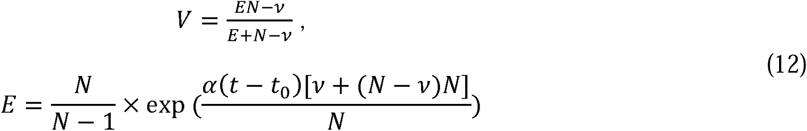

Finally, this approximation (Eq. (12)) reached a limited amount of victims, since *V = I + S* tends to N at infinity.

Figure 1 shows the prediction results of COVID-19 deaths in Iran based on the fitting of the available data of Iran to the epidemic curve of China in a mild scenario. Based on this scenario, 21.03.2020 would be the COVID-19 epidemic peak in Iran. After this point, the outbreak and death would be reduced so that this epidemic would be terminated at the end of April 2020.

**Fig. 1.**
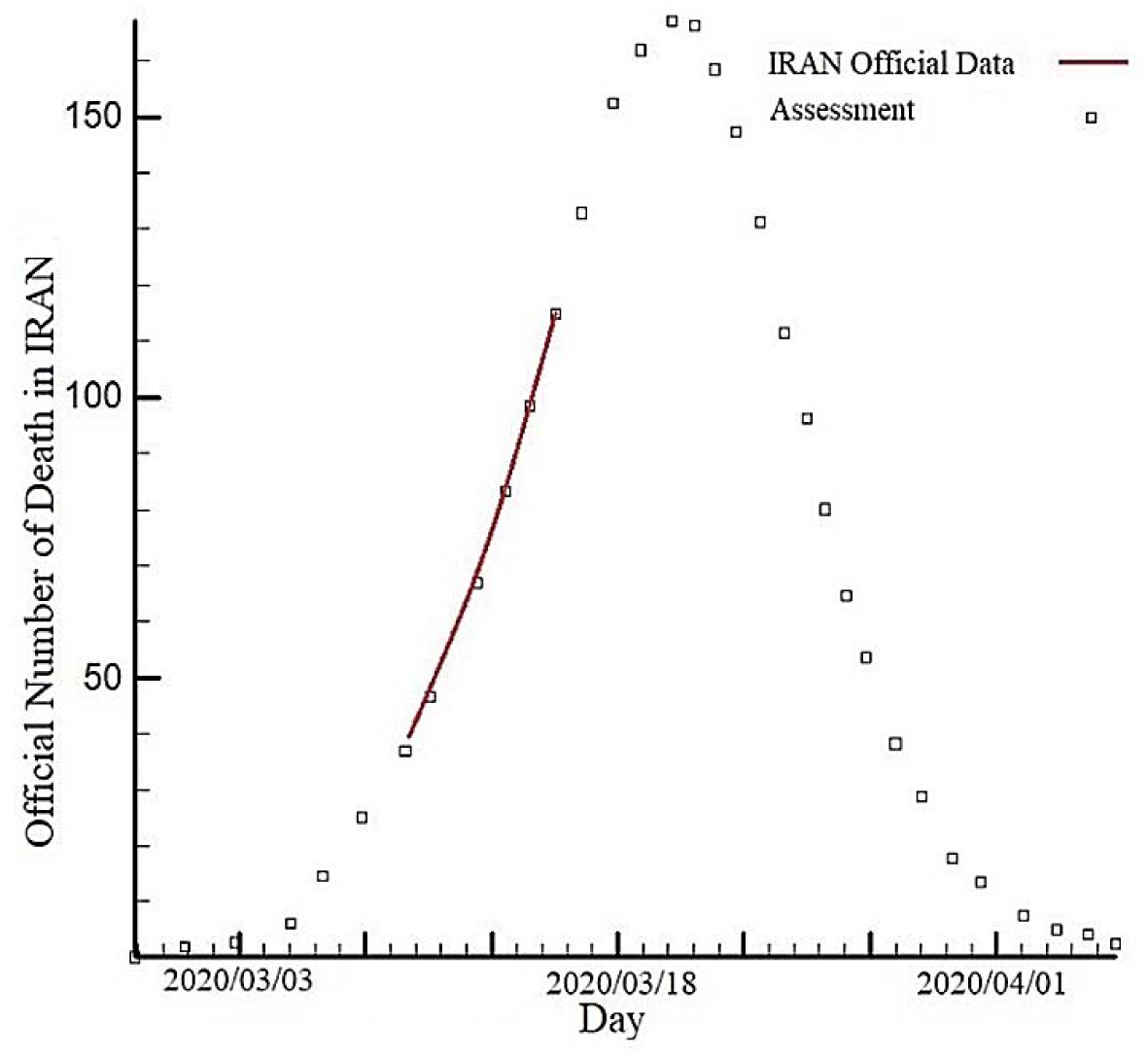
COVID-19 deaths in Iran based on the data of China optimistic perdition curve

The optimal calculated values of parameters are shown in Table. 1. Also, Table 2 lists the values obtained by the SIR model for the COVID-19 epidemic in Iran. As can be observed, the official statistical data of Iran for the susceptible and recovered people were used for the extrapolation process. It has been predicted that the maximum number of infected people would occur on 27.03.2020, and the disease peak would occur on 19.03.2020 (which was obtained based on the calculation of infection rate). Hence, due to a 2-to 3-day difference between death and infection peaks, there is a good agreement between the curves depicted in Figs. 1 and 2. The infection rate would be zero at the beginning of May 2020, and the number of susceptible people would converge to its lowest value.

**Table 1.**
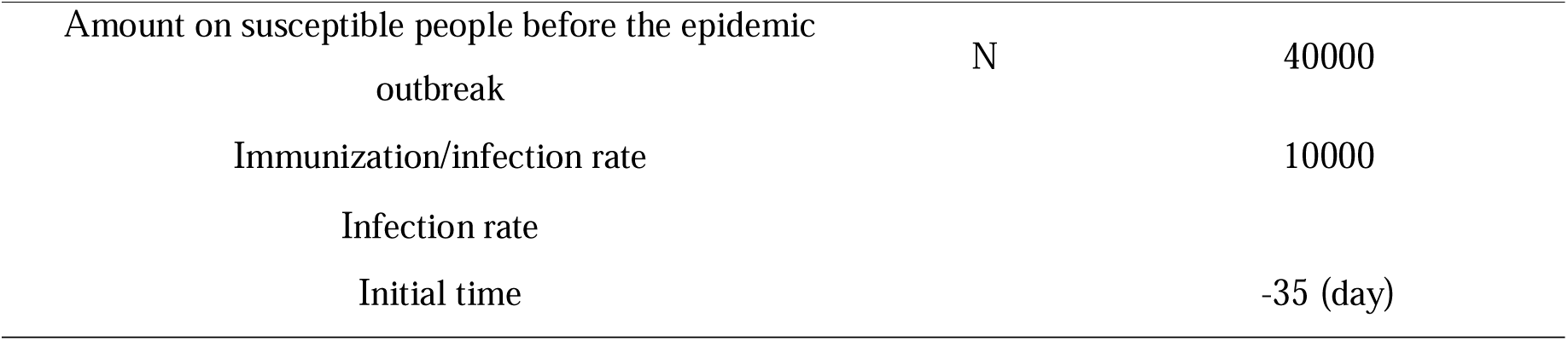
The optimistic optimal and values of parameters

|  |  |  |
| --- | --- | --- |
| Amount on susceptible people before the epidemic outbreak | N | 40000 |
| Immunization/infection rate |  | 10000 |
| Infection rate |  |  |
| Initial time |  | -35 (day) |

**Table 2.** The data of epidemic caused by the COVID-19 pneumonia in Iran [10].

| Date | Infection | Daily<br>Recovered | Death | Infection | Total<br>Recovered | Death |
| --- | --- | --- | --- | --- | --- | --- |
| 2020/2/19 | 2 | 0 | 2 | 2 | 0 | 2 |
|  | 3 | 0 | 0 | 5 | 0 | 2 |
|  | 13 | 0 | 2 | 18 | 0 | 4 |
|  | 11 | 0 | 2 | 29 | 0 | 6 |
|  | 14 | 1 | 2 | 43 | 1 | 8 |
| 2020/2/24 | 18 | 2 | 4 | 61 | 3 | 12 |
|  | 34 | 22 | 4 | 95 | 25 | 16 |
|  | 44 | 0 | 3 | 139 | 25 | 19 |
|  | 106 | 29 | 7 | 245 | 54 | 26 |
|  | 143 | 19 | 8 | 388 | 73 | 34 |
| 2020/2/29 | 205 | 50 | 9 | 593 | 123 | 43 |
|  | 385 | 52 | 11 | 978 | 175 | 54 |
|  | 523 | 116 | 12 | 1501 | 291 | 66 |
|  | 835 | 144 | 11 | 2336 | 435 | 77 |
|  | 586 | 117 | 15 | 2922 | 552 | 92 |
| 2020/3/5 | 591 | 187 | 15 | 3513 | 739 | 107 |
|  | 1234 | 174 | 17 | 4747 | 913 | 124 |
|  | 1076 | 756 | 21 | 5823 | 1669 | 145 |
|  | 743 | 465 | 49 | 6566 | 2134 | 194 |
|  | 595 | 260 | 43 | 7161 | 2394 | 237 |
| 2020/3/10 | 881 | 337 | 54 | 8042 | 2731 | 291 |
|  | 958 | 228 | 63 | 9000 | 2959 | 354 |
|  | 1075 | 317 | 75 | 10075 | 3276 | 429 |
|  | 1289 | 253 | 85 | 11364 | 3529 | 514 |
|  | 1365 | 810 | 97 | 12729 | 4339 | 611 |
| 2020/3/15 | 1209 | 251 | 113 | 13938 | 4590 | 724 |
|  | 1053 | 406 | 129 | 14991 | 4996 | 853 |
|  | 1178 | 393 | 135 | 16169 | 5389 | 988 |
|  | 1192 | 321 | 147 | 17361 | 5710 | 1135 |
|  | 1046 | 269 | 149 | 18407 | 5979 | 1284 |
| 2020/3/20 | 1237 | 766 | 149 | 19644 | 6745 | 1433 |
|  | 966 | 890 | 123 | 20610 | 7635 | 1556 |
|  | 1028 | 278 | 129 | 21638 | 7913 | 1685 |
|  | 1411 | 463 | 127 | 23049 | 8376 | 1812 |
| 2020/3/24 | 1762 | 537 | 123 | 24811 | 8913 | 1935 |

**Table 3.**
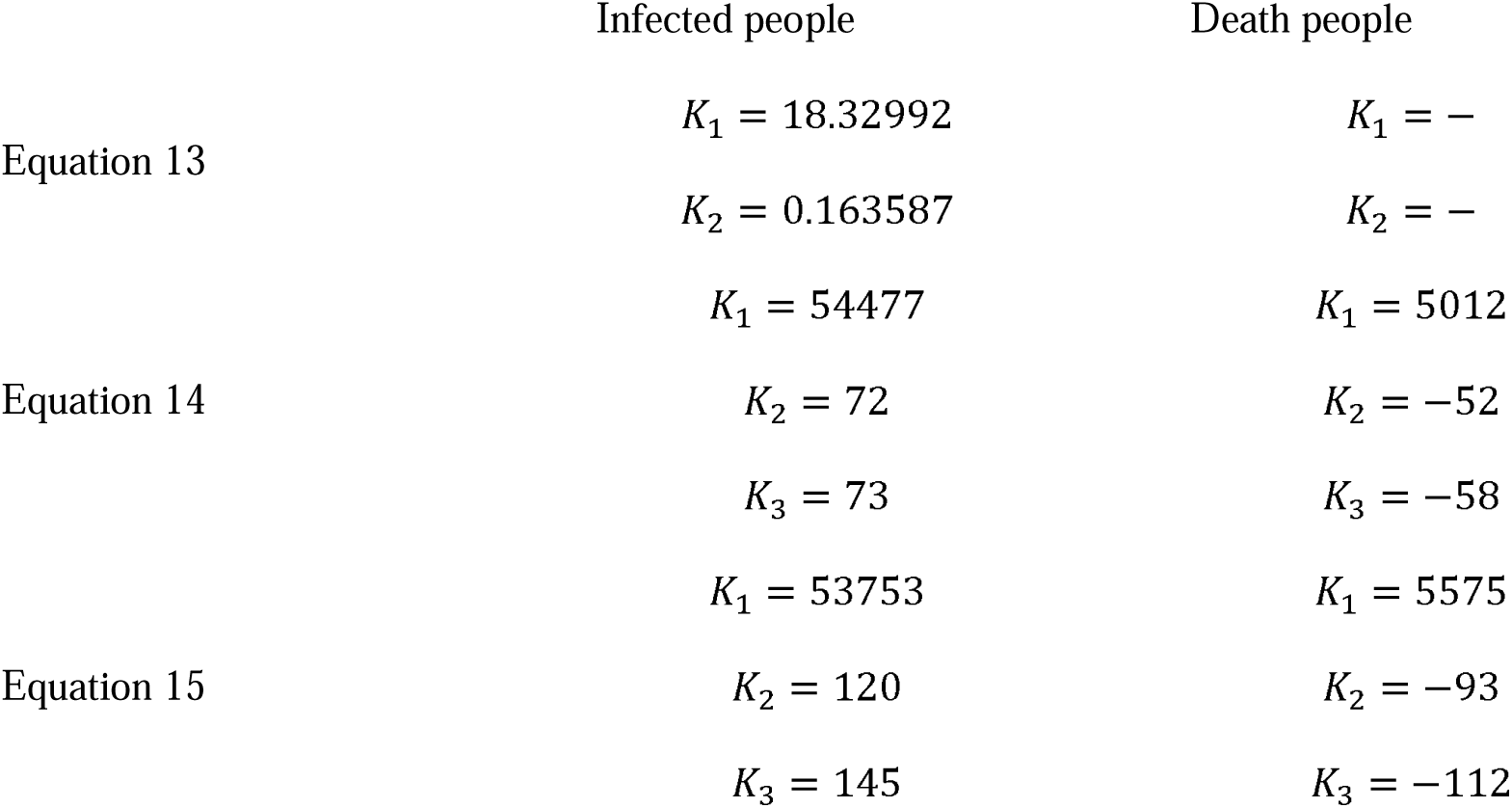
The constant value of extrapolation functions

**Fig. 2.**
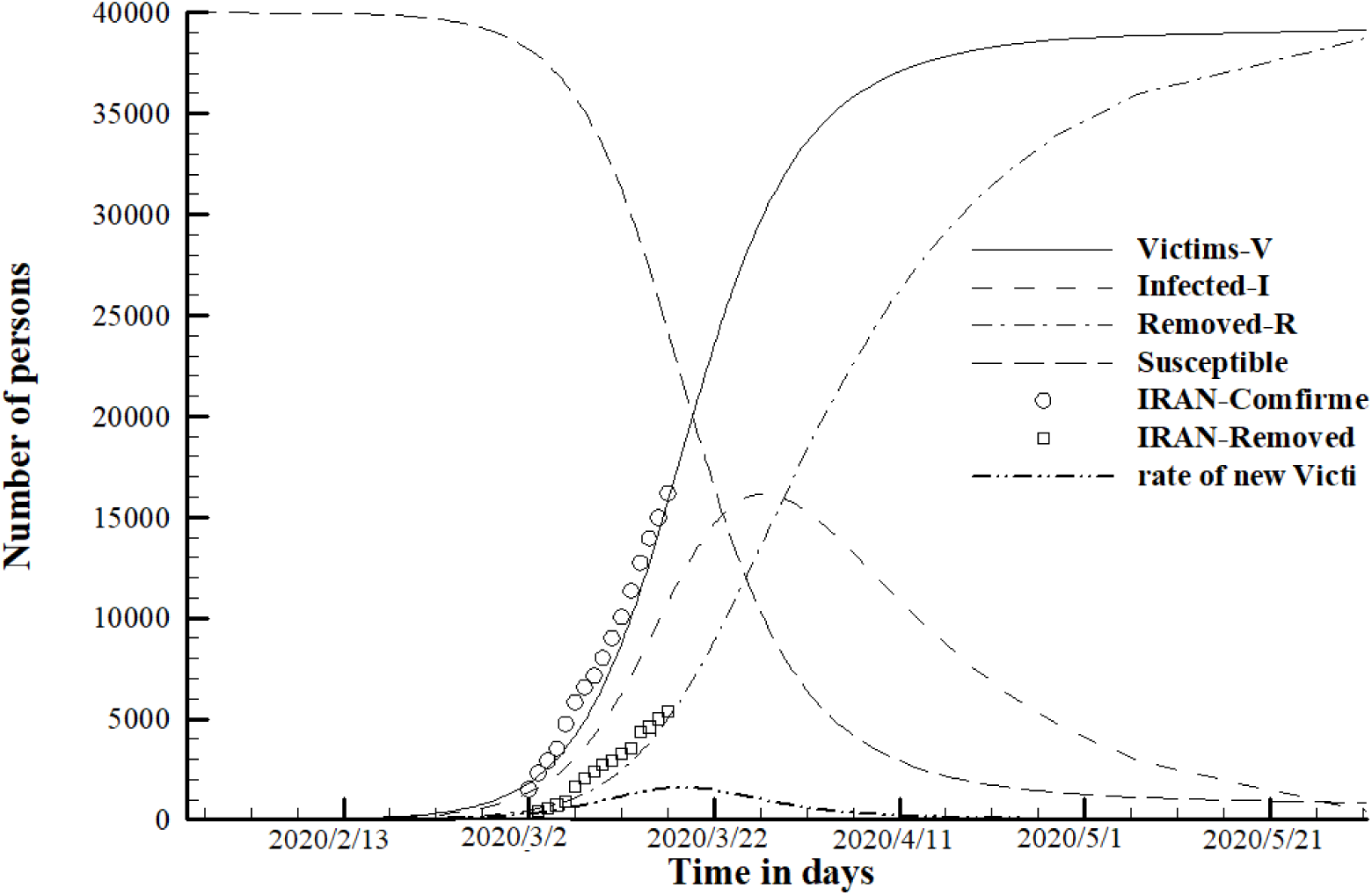
The values obtained by the SIR model for the COVID-19 epidemic in Iran

- STEP 2

In this section, we estimate the number of infected people by the disease in Iran with a total population of 84 million [14].

According to the articles on epidemics in China and Japan [4, 15], the proportion of people with the epidemic to the total population (*N/N_*total*_*) is as low as 1.89 to 3.58 percent. As this subject, here, we consider 2.14 percent. This means that the total number of people caught in Iran will be approximately 1.8 million. According to Figure 3, the number of people exposed to the disease will reach beside four hundred thousand but using a suitable diagnosis system and regarding hygiene behavior of susceptible persons and quarantine conditions, it is anticipated that disease spreading will be stopped and most of the victims will be recovered. As can be seen in Figure 4, the epidemic will continue by mid-June (late spring).

**Fig. 3.**
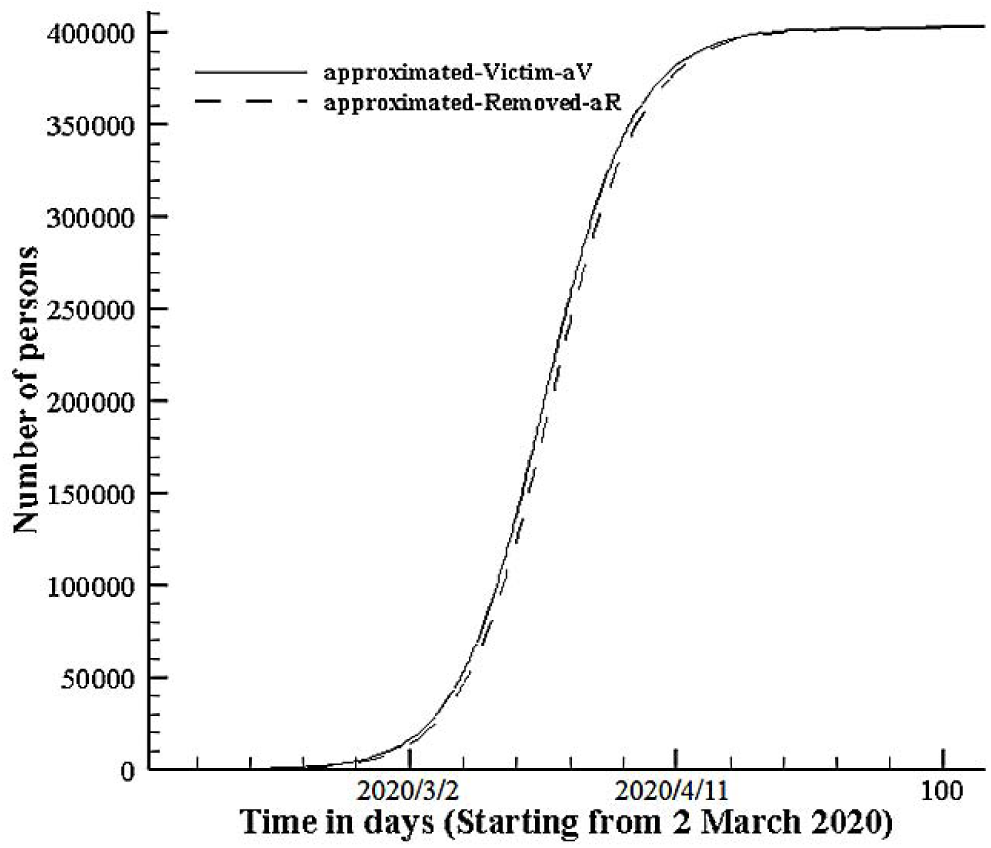
The approximation calculation of victim and removed people in Iran

**Fig. 4.**
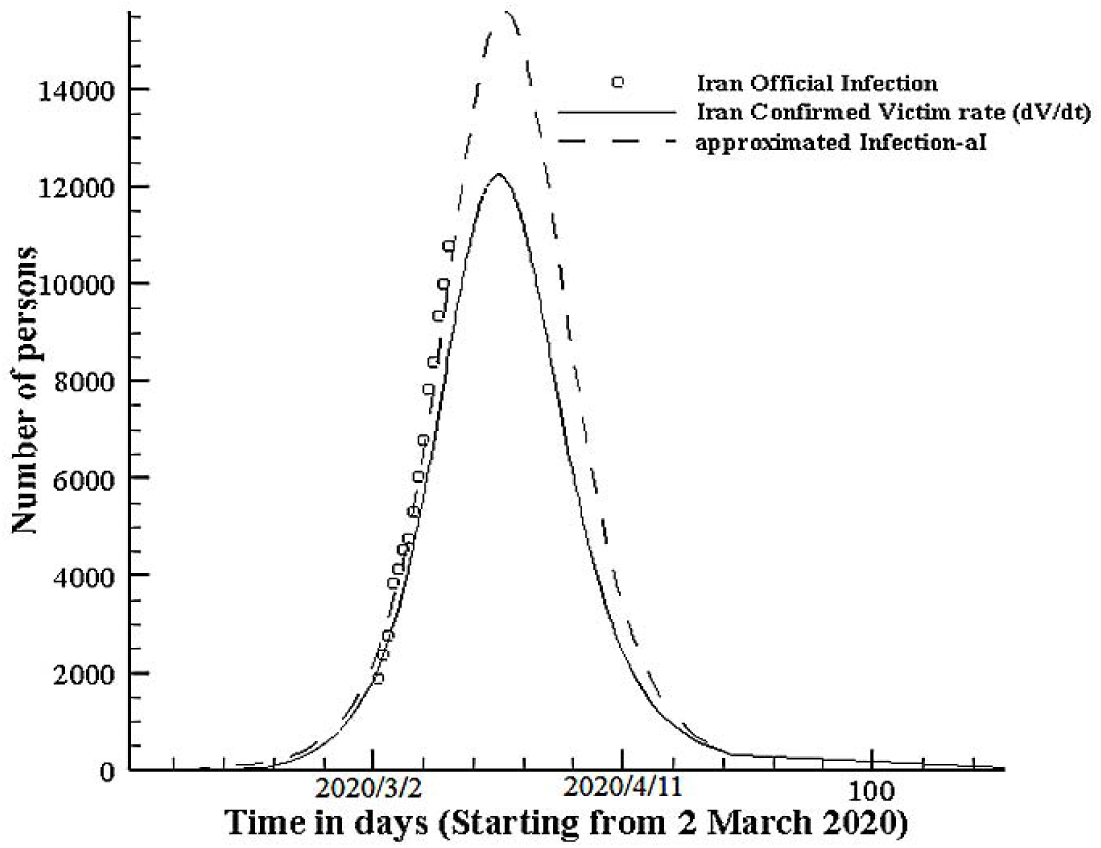
The number of approximated infection people and ultimate disease time in Iran

- STEP 3

In this step, statistical data of people who have caught the disease have been depicted from 19 February 2020 to 24 March 2020 in fig. 5 [10] and extrapolation was accomplished by using of least square method in the background of SIR model, moreover.

**Fig. 5.**
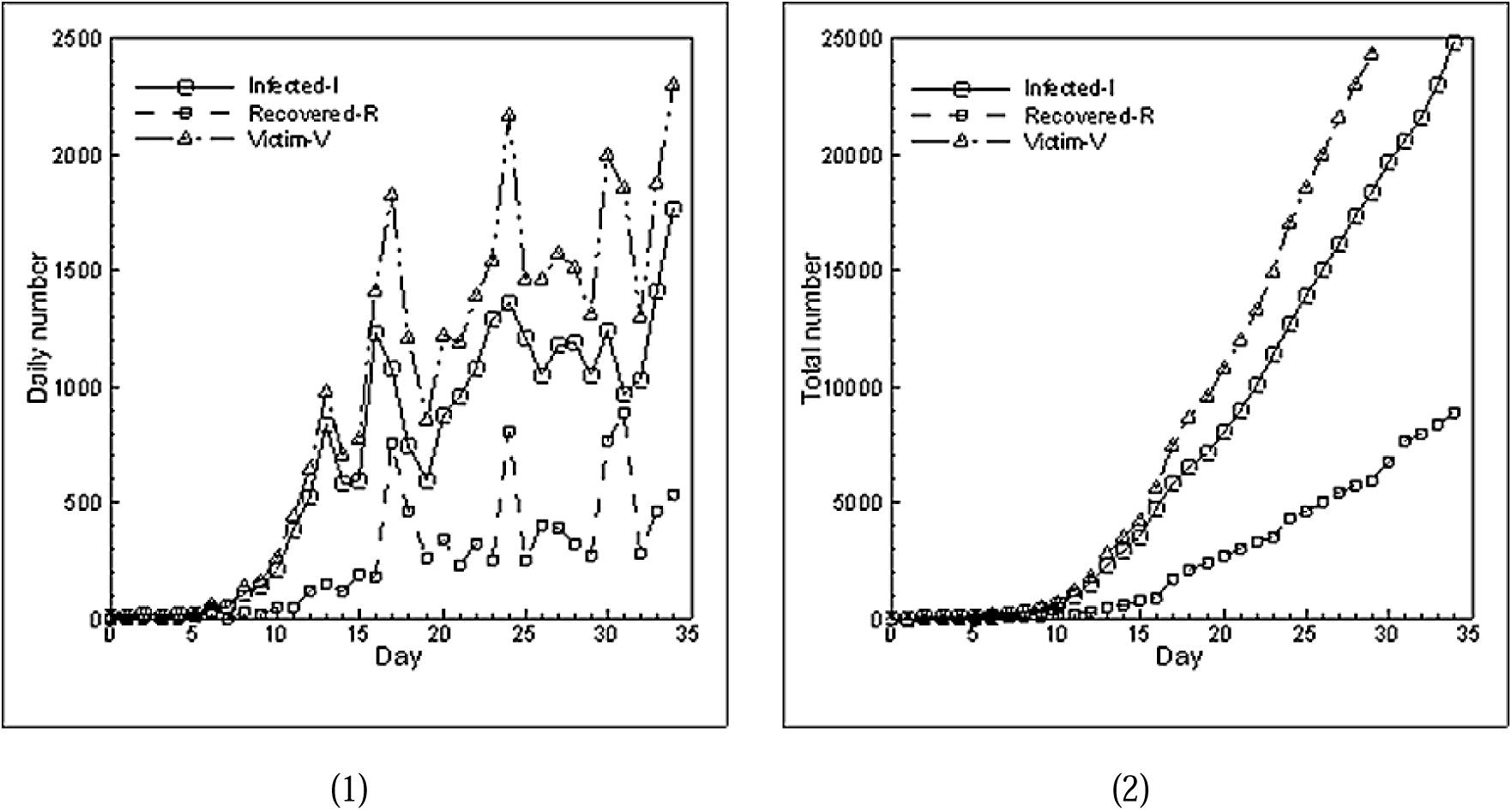
The number of Infected, Recovered, Victim people in Iran [10] (1) Daily (2) Total

**Fig. 6.**
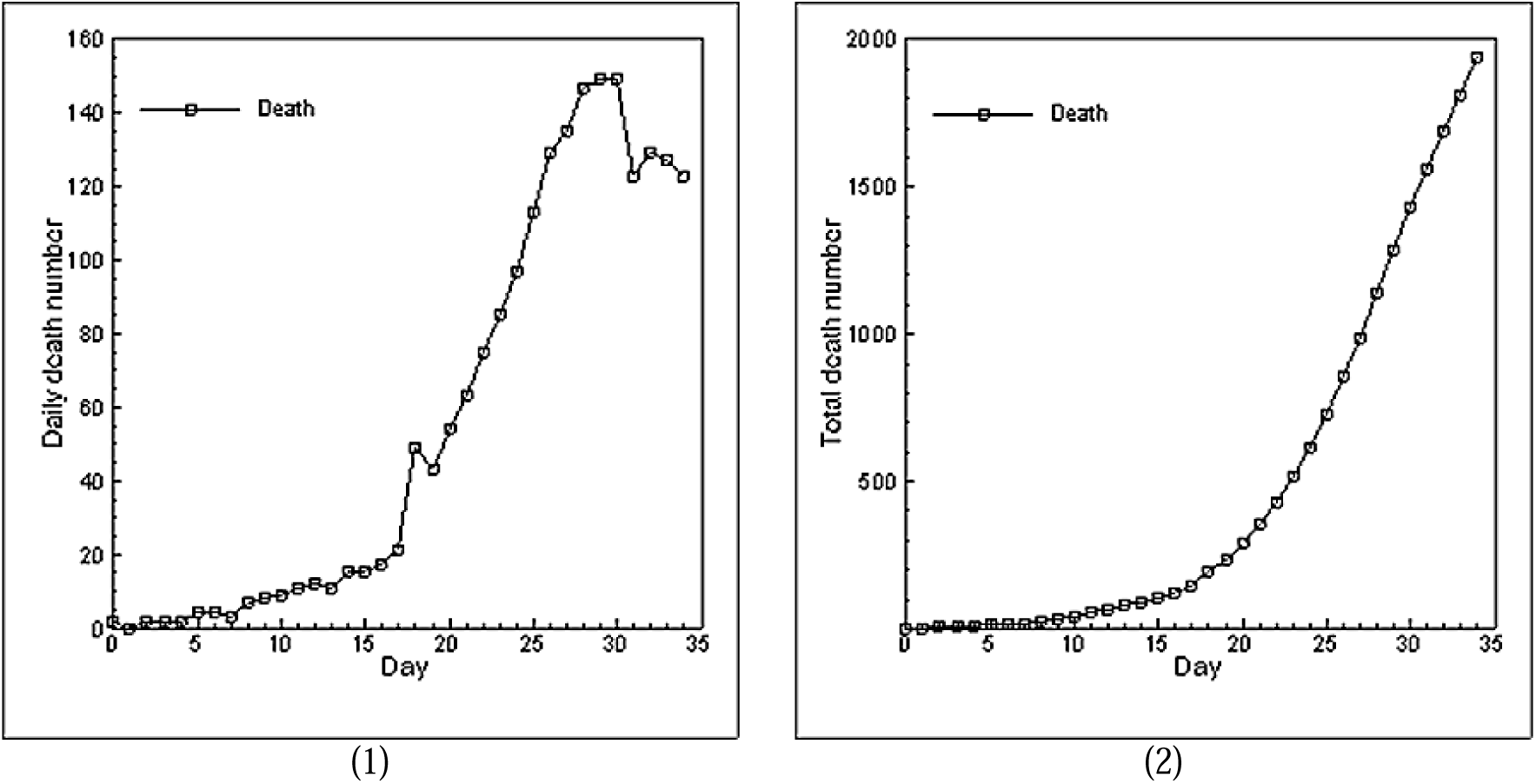
The number of death people in Iran (1) Daily (2) Total

This section deals with data extrapolation based on the least squares model. Three functions are used for extrapolation. The exponential function (equation 13) is the general function of the SIR methods. Periodic type functions (equations 14 and 15) is also used. The purpose of this is to consider if there is a possibility of multiple peaks until the epidemic ends [16]. As can be seen in Fig. 7, the data fits much closer with the general model of the exponential function (Equation 13) so it can be concluded that possibility to have different number of peaks to end of epidemic is negligible and exponential distribution function is the most appropriate model for forecasting the peak time.

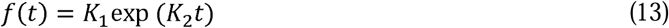

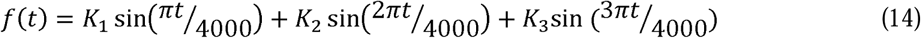

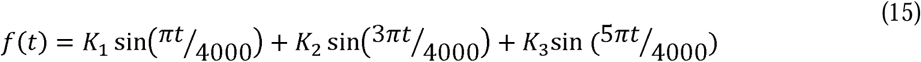

**Fig. 7.**
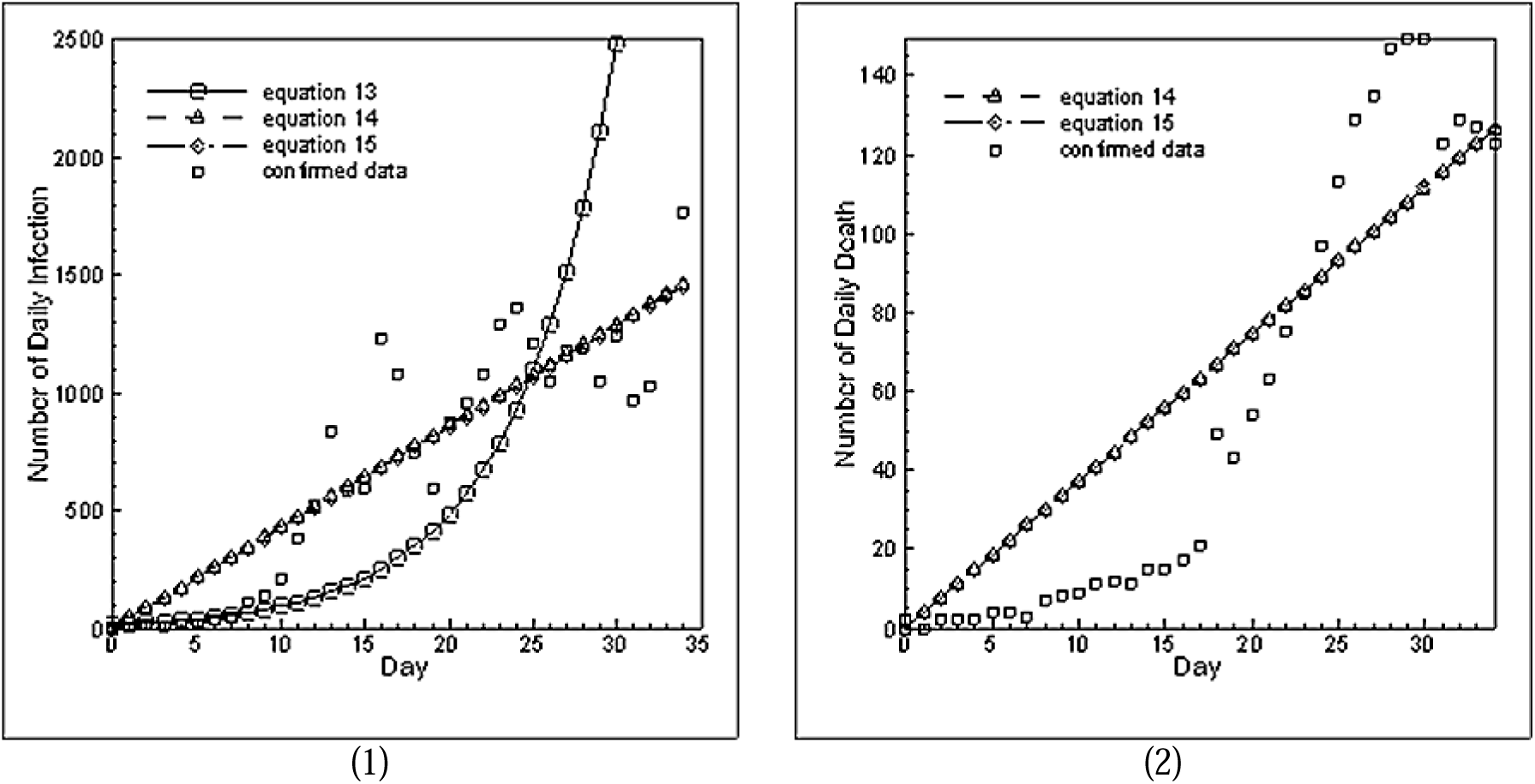
Different extrapolation functions (1) Daily Infections (2) Daily Death

In exponential formula t is known as time, calculated as *t* = *t*_*Day*_ − *t*_*Turning*_ and *t*_*Turning*_= 37 is achieved.

Ultimately, people can prevent from spreading the disease by doing some things such as home quarantine and personal hygiene [17, 18] since the infection rate has had low value, expecting a longer time with fewer infected and died people based on forecasting will be occur. Owning to, Coronavirus Incubation Period time to show symptoms is varied. This may appear 2-14 days (known more as 11.5 days) after exposure [19], it’s better to determine an interval time for peak time. According to this investigation, this interval is from 2020/3/27 to 2020/4/7. Hence, in this interval time, Covid-19 will be at its peak and the epidemic will disappear by mid-June.

## Concluding Remarks

In previous studies, the number of recovered people was not considered, which led to a significant error in the calculations. However, by incorporating the available official statistical values of recovered people along with the number of susceptible people in the present study, the accuracy of results obtained by the SIR model. Furthermore, another desirable feature of the current predictions is the fact that the estimation of susceptible people, who are still present in the population *S*_∞_ = *N* − *V*_∞_, has a suitably lower value compared with the previous works. It means that these people would not couch the infection after the suppression of the epidemic. Also, good agreement has been observed between the predictions obtained by fitting the official number of deaths (Fig. 1) and the results of the SIR model. By using the coefficient of the considered number to the total population, it is anticipated that the total number of infected people will reach four hundred thousand and finally, the epidemic will end in late spring.

## Data Availability

Available upon request

